# Machine Learning Improves the Predictive Utility of Lactic Acid in Hospitalized Infants

**DOI:** 10.1101/2025.04.01.25321818

**Authors:** K. Taylor Wild, Ibrahim George-Sankoh, Stephen R. Master, Rebecca D. Ganetzky

## Abstract

**Background and Objectives:** Hyperlactatemia is common in hospitalized infants. Machine learning was applied to clinical and laboratory characteristics in hospitalized infants with hyperlactatemia to identify predictors of inborn error of energy metabolism (IEEM).

**Methods:** Retrospective cohort study of hospitalized infants aged 0-90 days (2012-2020) with a lactate ≥ 5 mmol/L. Final diagnosis was discretized to IEEM, cardiac, hypoxia, infectious and other. Random forest and XGBoost models were tuned and compared using cross-validation, and a final model was evaluated on an independent test set to determine ability to predict IEEM.

**Results:** Among 1000 infants, median lactate was 8 mmol/L. The overall mortality rate was 30%, (N=291) and was 51% (N=21) among infants with an IEEM (N=41). Lactate was significantly higher in infants with an IEEM (12.6 mmol/L; IQR: 5-27 mmol/L). Machine learning analysis including plasma amino acid and acylcarnitine values yielded an area under the ROC curve (AUC-ROC) of 0.81 in a held-out test set, and was significantly better than lactate alone in a comparable population (AUC-ROC 0.81 vs. 0.56, p=0.027).

**Conclusions:** Rapid diagnosis of IEEM vs. other causes is essential for neonatal hyperlactatemia prognostication. Machine learning has high diagnostic utility, serving as a framework for computer-aided interpretation of complex diagnostic data.

## Introduction

Lactic acidosis is a relatively common finding in hospitalized infants. As a byproduct of anaerobic metabolism, lactate can serve as a surrogate maker of inadequate tissue oxygenation. Neonatal lactic acidosis may be due to an inborn error of energy metabolism (IEEM), which is also known as primary lactic acidosis. Hyperlactatemia can also be secondary in the setting of cardiac dysfunction, tissue anoxia, and severe catabolic states, such as infection and severe dehydration. Common neonatal conditions that may result in poor tissue oxygenation include many forms of congenital heart disease (CHD), hypoxic ischemic encephalopathy (HIE), meconium aspiration syndrome (MAS), persistent pulmonary hypertension of the newborn (PPHN), and congenital diaphragmatic hernia, among various other diagnoses. In neonates, hyperlactatemia may also be spurious as lactate is extremely sensitive to sample collection technique and handling.^1,2^

In neonates with congenital heart disease (CHD), peak lactate has been shown to be predictive of early morbidity and mortality, as well as post-surgical residual lesions that may prompt earlier investigation and intervention.^3^ Similarly, in neonates with HIE, Shah et al. found that significant hyperlactatemia was associated with moderate-or-severe HIE with a sensitivity of 94% and specificity of 67%. They also found that initial lactate levels were significantly higher (p=0.001) in neonates with moderate-to-severe HIE (mean +/-SD=11.09+/-4.6 mEq/L) as compared to those with mild or no HIE (mean +/-SD=7.1+/-4.7 mEq/L). Lactate levels remained elevated longer in the infants with moderate-to-severe disease.^4^

IEEM that cause lactic acidosis (primary lactic acidosis) include 1) electron transport chain dysfunction 2) disorders of pyruvate metabolism or the tricarboxylic acid cycle, when pyruvate cannot enter the citric acid cycle and is shunted back into lactate, 3) organic acidurias, in which accumulating organic acids are toxic to mitochondrial energy production and also compete with lactate for urinary excretion, 4) fatty acid oxidation defects due to secondary impairment of the electron transport chain and 5) gluconeogenic defects. Overall, primary lactic acidosis most often occurs due to genetic defects in the electron transport chain or disorders of pyruvate metabolism, whereas secondary lactic acidosis can occur from many different causes of impaired tissue oxygenation.^2,5-7^

The association between neonatal hyperlactatemia and IEEM has been less well-described. Mitochondrial dysfunction may present in the perinatal period because of the high energy requirements of the growing newborn. While neonates with mitochondrial disease often have a primary lactic acidosis as their predominant phenotype, they may be difficult to differentiate from other critically ill infants. Presenting symptoms may be non-specific and highly variable, including hypotonia, respiratory distress, apnea, feeding difficulties, failure to thrive, and seizures.^8^ Few studies have investigated large groups of neonates with mitochondrial disease.^5-7^ Therefore, it is unclear how many neonates with hyperlactatemia have an underlying genetic mitochondrial dysfunction compared to secondary causes of hyperlactatemia.

This study aimed to analyze the clinical and laboratory characteristics of all infants hospitalized at the Children’s Hospital of Philadelphia between 0-90 days of life found to have hyperlactatemia, assisted by a machine-learning approach to identify laboratory signs and phenotypic features predictive of primary hyperlactatemia. Hyperlactatemia was defined as a lactate level greater than or equal to 5 mmol/L to evaluate infants with severe hyperlactatemia and with an increased risk of an IEEM above this level.^2,5,6^

## Methods and Materials

A retrospective chart review was performed on all infants 0-90 days of life admitted to the Children’s Hospital of Philadelphia from January 2012 to August 2020 with a lactate level greater than or equal to 5 mmol/L. This study was deemed exempt by the Institutional Review Board at the Children’s Hospital of Philadelphia. Automated data extraction was performed using the Alteryx platform. For each infant data collected were: age, sex, genetic testing, echocardiogram findings, need for extracorporeal membrane oxygenation, age at death, plasma amino acid analysis, plasma acylcarnitine analysis, urine organic acids, height of peak lactate and age at peak lactate. Based on the listed final diagnosis at discharge, patients were coded as CHD, IEEM, HIE, sepsis/serious infection, or other. Echocardiogram reports were manually reviewed by a physician. When available, the first echocardiogram and the echocardiogram closest to the time of the peak lactate were reviewed. Patients were coded for presence or absence of pulmonary hypertension, diminished cardiac function, cardiomyopathy, or structural heart disease. Metabolic testing was reviewed manually by geneticists. When multiple values were available, the first levels drawn were selected. Amino acids, acylcarnitines, pyruvate levels, and pyruvate/lactate ratios were analyzed as continuous variables. Absolute values were reported for continuous variables. Because urine organic acids over the course of this study were analyzed on a semi-quantiative platform with only final interpretations available in the electronic medical record, these were analyzed as a categorical variable, with a determination of normal/abnormal was made based on the test interpretation by the interpreting metabolic laboratory director, all of whom were either board-certified clinical biochemical geneticists or a board-certified clinical chemist. The presence lactaturia was expected based on the degree of lactatemia and was not considered in the classification of organic acids.

Of the 1000 cases, full data including quantitative plasma amino acids and plasma acylcarntine levels was available for 150 subjects. Machine learning was performed in R version 4.4.1^9^ using the tidymodels, ranger, and xgboost packages.^10-12^ The 150 cases with amino acid and acylcarnitine data were randomly partitioned into a training (n=112) and test (n=38) set. Machine learning models (Random forest tuned vs. untuned, XGBoost tuned vs. untuned) were optimized and compared using cross-validation on the 112-sample training set. Bagging was used to impute missing values, redundant correlated variables (<0.9) were filtered, and zero-variance variables were removed. Based on cross-validation performance in the training set, the tuned XGBoost model was chosen as the optimal method (S. Table 1) and trained on the 112-sample set. The full trained model was validated using the held-out 38-sample test set. Global variable importance was assessed using the vip package.^13^ [Greenwell and Boehmke, 2020], and additional local variable importance was assessed using SHAP analysis of the training set using the SHAPforxgboost package.^14^ [Liu and Just, 2023]. R code and deidentified data necessary to reproduce our reported results are provided as a supplementary file [S. File 1].

Survival was calculated by Kaplan Meier curve analysis with Cox proportional hazards regression using the R packages survival and survminer.^15-17^ Statistical comparison of ROC curves was performed by DeLong’s test using the pROC package.^18^

For multiple comparisons analysis of variance (ANOVA) using a Tukey honestly significant difference (HSD) was performed. Values were considered statistically significant if the p-value was less than 0.05.

## Results

Cohort characteristics are shown in Table 1. There were 1,000 infants between 0-90 days of life with at least one lactate level greater than or equal to 5 mmol/L. The median peak lactate level was 8 mmol/L (interquartile range (IQR) 6, 12). There were 59% males (N=585) and 41% females (N=415). Echocardiograms were available and reviewed for 842 infants. Full metabolic profile (amino acids, acylcarnitine profile and pyruvate/lactate) was available for 114 cases.

**Table 1:**
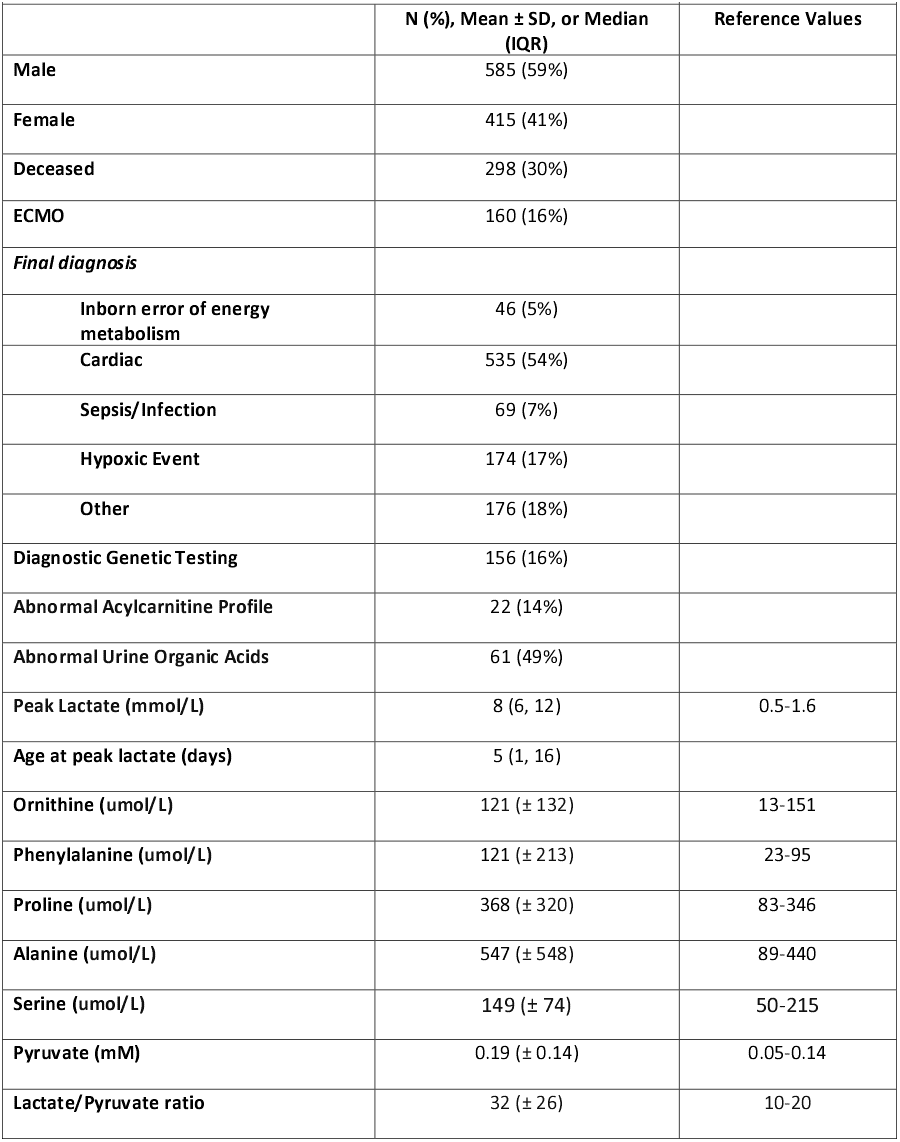
Cohort Characteristics (N=1,000)

CHD was the most common cause of hyperlactatemia during the first 90 days of life (N=535, 54%), followed by hypoxic ischemic encephalopathy (HIE) (N=174, 17%), sepsis/infection (N=69, 7%), IEEM (N=46, 5%), or “Other” for a diagnosis that did not fit into one of those primary categories (N=176, 18%). Among some of the most common “Other” causes were congenital diaphragmatic hernia (N=22, 2%), neuroblastoma (N=4, 0.4%), meconium aspiration without HIE (N=4, 0.4%), and autosomal recessive polycystic kidney disease (N=4, 0.4%).

Lactate levels varied by primary diagnostic category (p=0.002, ANOVA; Figure 1). Lactate was significantly higher in infants with an IEEM (median 11.6 mmol/L, IQR 8,15) compared to infants with CHD, HIE or other causes of lactic acidosis (IEEM vs CHD (p=0.01, ANOVA), IEEM vs HIE (p=0.02, ANOVA), and IEEM vs other (p = 0.001, ANOVA) (FIG 1A). Lactate also significantly predicted survival status, with an average peak lactate of 12.5mmol/L in patients who ultimately passed away and 8.7mmol/L in survivors, (p<2.2*10^−16^) as well as showing differences between survivors and decedants in all subcohorts. (FIG 1A) The most common types of CHD were hypoplastic left heart syndrome (N=141), transposition of the great arteries (N=65), tetralogy of fallot (N=46), coarctation of the aorta (N=42), truncus arteriosus (N=29), common AV canal (N=30), double outlet right ventricle (N=28), and pulmonary atresia (N=25). Among all 842 infants with echocardiograms, common findings included evidence of diminished function (N=211), pulmonary hypertension (N=191), and evidence of cardiomyopathy (N=41) based on echocardiogram report.

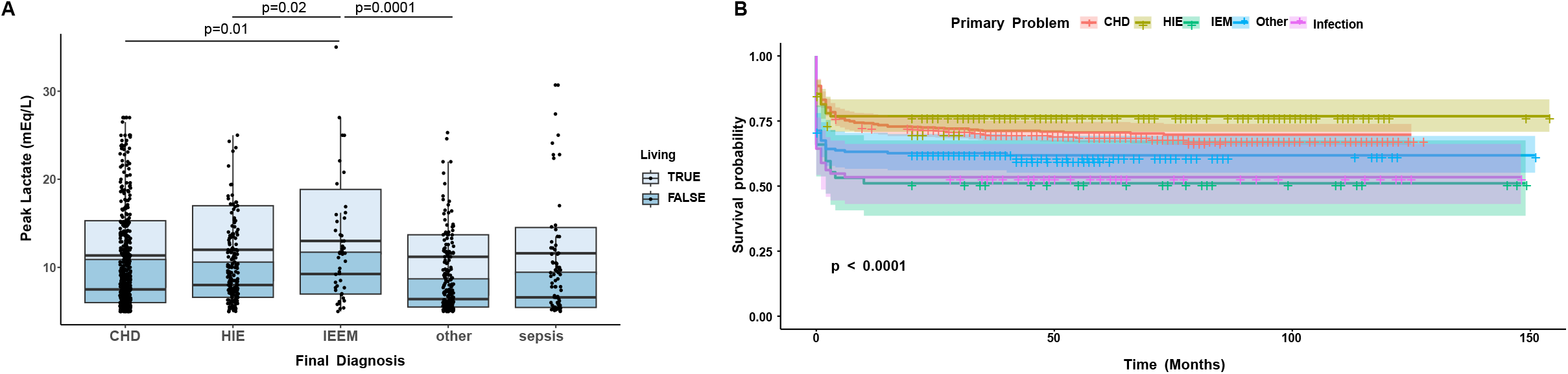

The mortality rate was 34% (N=338) overall, but significantly higher (54% (N=25)) among infants found to have an IEEM. Final diagnosis correlated with mortality (p<0.0001), with significantly higher mortality in the IEEM compared to the HIE cohort (p=0.004) and CHD cohort (p=0.002). (FIG 1B) Infants who survived to 20 months of age were likely to survive long-term, regardless of underlying cause. Extracorporeal membrane oxygenation (ECMO) was required in 16% of infants (N=160). Genetic testing, predominantly exome and genome sequencing, was diagnostic in 16% of all infants (N=156), including 91% (N=42) of infants with a clinical and laboratory diagnosis of IEEM. Specific diagnoses included long chain fatty acid oxidation defects (N=9) [most commonly, trifunctional protein deficiency (N=6)] organic acidurias [including methylmalonic acidemia (N=4), cobalamin C & glutaric aciduria], urea cycle disorders (N=3), glycogen storage disease (N=3) [including glycogen storage disease 1b (N=2)], pyruvate kinase deficiency, pyruvate dehydrogenase deficiency, pyruvate carboxylase deficiency, citrate transporter deficiency, primary mitochondrial respiratory chain disease associated with variants in *TRMU* (n=3), *SLC25A3, FBLX4, NDUFB11, FOXRED1, ECSH1, YARS2, FARS2, POLG*, single large scale mtDNA deletion & *ATAD3* regional copy number variation.

Plasma amino acids were available for 172 infants (Table 1). Pyruvate levels were available for 127 infants with a median of 0.16 mM (IQR 0.09, 0.26). Lactate/Pyruvate ratios were available for 126 infants with a median of 25 (IQR 17, 37.75). Acylcarnitine profiles were available for 183 infants and were abnormal in 14% (N=22) of infants. Urine organic acids were available for 125 infants and were abnormal in 49% (N=61) of infants.

Given the prognostic significance of IEEM, we next explored whether it was possible to classify patients as IEEM using laboratory test results. We therefore created and validated a machine learning classifier to perform this assessment. To ensure that adequate numbers of samples were available for training and testing sets, analysis included cases with plasma amino acid and acylcarnitine results (N=150) including some in which the pyruvate/lactate ratio was not measured. The variables used to develop the classifier were sex, peak lactate, age at presentation, and quantitative plasma amino acid and acylcarnitine values. Correlation between variables in this cohort was visualized (supplementary figure 1). Reported race was analyzed as a descriptive variable to ensure equitable representation of the data set but was not included in the classifier. Cross-validation on the training set (N=112) was used to compare algorithms (random forest vs. XGBoost, untuned vs. tuned) using area under the receiver operating characteristic (ROC) curve (AUC-ROC) as the target metric. Results of the cross-validation are shown in supplementary table 1, and a tuned gradient boosting (XGBoost) model was chosen as the best model. This algorithm was then retrained using the full training set, and model performance was assessed on the independent test set (supplementary table 2).

The full trained XGBoost model had an accuracy of 89.5% on the test set, an area under the receiver-operator characteristic curve of 0.81 (FIG. 2A), and an area under the precision-recall curve of 0.94. In contrast, a ROC curve reflecting the performance of peak lactate alone in all cases (N=1000) yielded an AUC of 0.65 (Supplementary Figure 2). To ensure that analysis was being performed on comparable subpopulations, an ROC curve for lactate was also generated using only cases for which amino acid and acylcarnitine results were available (N=150), corresponding the training+test set used for machine learning. In this population, lactate alone had an area under the receiver-operator characteristic curve of 0.56 (Fig. 2A), and the difference between these lactate-only and machine learning ROC curves in comparable populations (0.56 vs. 0.81) is statistically significant (p = 0.027).

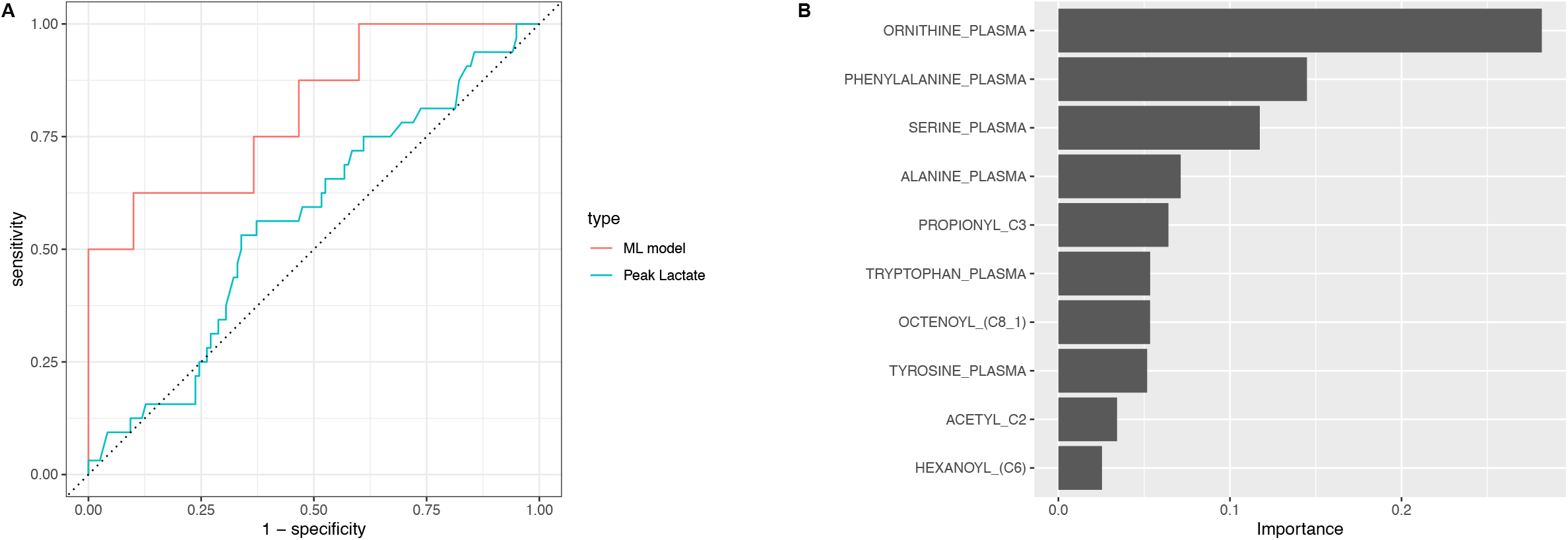

We then assessed global variable importance of our machine learning model. The variables of highest importance included plasma levels of the amino acids ornithine, phenylalanine and serine in addition to the expected variables such as height of peak lactate, pyruvate, alanine and proline levels (FIG 2B). SHAP analysis (local variable importance) of the training showed similar results, (Supplementary Fig 3A), and initial analysis of SHAP patterns for training set samples shows multiple patterns of analytes contributing to the prediction results (Supplementary Fig 3B). Confirmatory univariate analysis demonstrated that serine levels were significantly higher in patients with IEEM compared to other diagnoses (Supplementary Fig 4).

## Discussion

The association between neonatal hyperlactatemia and IEEM is poorly understood. Because hyperlactatemia can also be secondary to other relatively common neonatal conditions such as cardiac dysfunction, tissue anoxia, and severe catabolic states, it is unclear how many neonates with hyperlactatemia are found to have an underlying genetic mitochondrial dysfunction compared to secondary causes of hyperlactatemia. This study aimed to identify laboratory signs and phenotypic features predictive of primary hyperlactatemia.

We found that the magnitude of the lactate peak was significantly higher among infants found to have an IEEM; however, this alone did not discriminate infants with IEEM from other causes of lactic acidosis. Infants with other causes of poor tissue oxygenation, particularly CHD, HIE, and sepsis, were often found to have significant hyperlactatemia. In particular, although as a group, lactate was much lower in infants with sepsis, individual patients with sepsis had the highest lactate levels in the cohort. As a surrogate marker of inadequate tissue perfusion, the height of lactate elevation is significantly associated with increased risk of death. As unbiased genetic testing was diagnostic in 15% of the overall cohort and in 90% of infants who had laboratory criteria consistent with an IEEM, expedited comprehensive genetic testing, such as rapid exome or genome sequencing, should be utilized in infants with profound hyperlactatemia of unclear etiology. This is particularly important given that mortality significantly varies by underlying cause with 30% mortality in any infant with hyperlactatemia compared to 51% mortality in infants with an IEEM. Therefore, early accurate diagnosis of IEEM is important for prognostication.

Similar to studies by Valencia et al. and Shah et al.^3-4^, we found marked hyperlactatemia in our infants with CHD and HIE, respectively; however, hyperlactatemia is significantly higher in infants with laboratory criteria consistent with an IEEM. In 2008, Gibson et al. found a diagnostic genetic result in 11/107 infants with an IEEM.^7^ Our diagnostic rate is significantly higher and likely reflects the significant advances that have occurred in molecular diagnostics. We anticipate that this rate will continue to increase as comprehensive genetic testing such as exome and genome sequencing becomes more readily available.

While expedited comprehensive genetic testing is of high diagnostic yield, clinical and metabolic testing remain faster. In our center with an in-house metabolic laboratory, testing for the most contributory metabolites can be completed in under 24 hours, including on weekends. In addition, metabolic testing is a critical component to helping validate genetic testing of uncertain significance and identifying patients who are missed by genetic testing. In this study ∼10% of infants who were ultimately diagnosed with an IEEM did not have diagnostic genetic testing in the neonatal period. Clinically and biochemically, there is no single variable that differentiates an infant with IEEM from CHD, sepsis, or HIE, etc.; however, using a multivariate machine learning classifier we were able to attain a diagnostic accuracy of 89.5%. This classifier performs significantly better than lactate alone, and in our opinion better than an unaided expert clinical diagnosis. Unexpectedly, the strongest predictors of an IEEM in our model were ornithine level, phenylalanine level, and serine level. Serine is an intriguing potential biomarker of IEEM in infants with hyperlactatemia. Serine levels in skeletal and heart muscle have previously been seen in mouse models of mitochondrial deficiency.^19^ Mechanistically, glucose is converted to serine through an enzyme phosphoglycerate dehydrogenase, which is a regulated step based on the NADH/NAD^+^ balance. Therefore, patients with mitochondrial disease, who are unable to oxidize NADH may have increased serine production. Elevated plasma serine has not been previously observed in patients with mitochondrial disease; however, by selecting for patients with profound lactic acidosis, this cohort may be enriched with a subgroup of patients with mitochondrial disease who have extreme elevation of NADH/NAD^+^. Given this role in energy production and maintenance, in this unique subpopulation, plasma serine may be a valuable biomarker. The role of ornithine and phenylalanine is unknown; these may reflect liver dysfunction, but other markers of liver dysfunction were not seen. Therefore these may reflect other, yet unknown, variables.

Limitations of this study include that is that it was a retrospective chart review and therefore a non-randomized sample. This study was limited by the existence of an electronic medical record, which began consistently around 2012. The use of exome sequencing has also increased significantly throughout the years of this study, so the rate of diagnostic genetic testing may be even higher in an era of advanced diagnostics. It is also possible that other variables that were not analyzed, such as other chemistry labs, the area under the lactate curve, etc. may also be important distinguishers. However, because of the size and heterogeneity of the sample as well as differences in practice over time, the number of variables that could be considered was limited. A major limitation is that only a minority of subjects included in this study had metabolic testing, thus there may have been infants with other causes of lactic acidosis (CHD, sepsis, etc), who may have an underlying IEEM that was not fully investigated. We hypothesize that the patients for whom there was full metabolic testing are those where the diagnostic possibility of an IEEM was higher on the differential, which is a potential bias in the data.

However, we feel that this is also a strength: that by developing an algorithm that is most skilled at looking at cases of high diagnostic uncertainty, we have maximized its utility for the real-life clinic setting. We also hope that these data, showing the value of metabolic laboratory data in achieving a definitive diagnosis, encourage more metabolic testing in this patient population as a low-cost, high turn-around time complement to genomic sequencing.

There were multiple strengths of our study. The first is that this is the largest published cohort of neonates with lactatemia from a single quaternary pediatric hospital. Our referral center included a wide spectrum of diagnoses with a large cardiac intensive care unit and neonatal intensive care unit. Detailed genetic and biochemical testing and echocardiograms were all individually evaluated for each patient. This was the first study to apply statistical learning to lactatemia in any patient population. Lactatemia is frequently clinically confounding – its presence indicates serious illness, but leaves few clues about the underlying cause. By applying supervised machine learning analysis, we have discovered novel predictors to aid in the diagnosis of the neonate with lactatemia. We believe that our results provide a strong rationale for a prospective study in which plasma amino acid and acylcarnitine data are collected more broadly on patents with lactatemia in order to validate their utility in rapid identification of IEEM.

## Conclusion

Lactatemia is a relatively common finding among hospitalized neonates. Lactate levels are statistically higher in neonates with an IEEM, but do not fully differentiate this group from neonates with lactate elevated secondary to other conditions. Metabolic screening labs and comprehensive genetic testing, such as rapid exome or genome sequencing, have a role in determining the underlying diagnosis of infants with persistent hyperlactatemia. Genetic testing is essential for definitive diagnosis. Serine may serve as a novel biomarker of primary genetic mitochondrial dysfunction. Accurate prediction is possible from clinical phenotyping and biochemical screening, allowing for the most rapid diagnosis. It is important to identify the etiology of hyperlactatemia as soon as possible because a diagnosis of primary IEEM has serious prognostic implications.

## Data Availability

The data that support the findings of this study are available as part of the supplementary distribution file included with this manuscript.

## Abbreviations

IEEM: Inborn Error of Energy Metabolism
HIE: Hypoxic Ischemic Encephalopathy
MAS: Meconium Aspiration Syndrome
PPHN: Persistent Pulmonary Hypertension of the Newborn
CHD: Congenital Heart Disease
ROC: Receiver Operator Characteristic
AUC: Area Under the Curve
PR: Precision Recall XGB: XGBoost
RF: Random Forest
IQR: Interquartile range
ANOVA: analysis of variance
HSD: honestly significant difference

## Data Availability

The data that support the findings of this study are available upon request.

## Acknowledgements

The authors would like to thank Laura MacMullen, who helped with submission of this protocol for IRB Exempt status

## Author Contributions

KTW, SRM, and RDG conceptualized and designed the study, collected and analyzed the data, drafted the initial manuscript, and reviewed and revised the manuscript.

IG-S assisted in collecting and analyzing the data and reviewed and revised the manuscript. All authors approved the final manuscript as submitted and agree to be accountable for all aspects of the work.

## Ethics Declaration

The CHOP Institutional Review Board granted this study exempt status.

## Conflict of Interest/Disclosures (includes financial disclosures)

Rebecca Ganetzky receives consulting fees from Minovia Therapeutics and is a paid advisor for Nurture Genomics. Stephen Master is a member of advisory boards for Indigo BioAutomation and Roche Diagnostics. The other authors have no conflicts of interest to disclose.

