## Supplementary figures and images for "Machine Learning Improves the Predictive Utility of Lactic Acid in Hospitalized Infants"

### S Fig 1

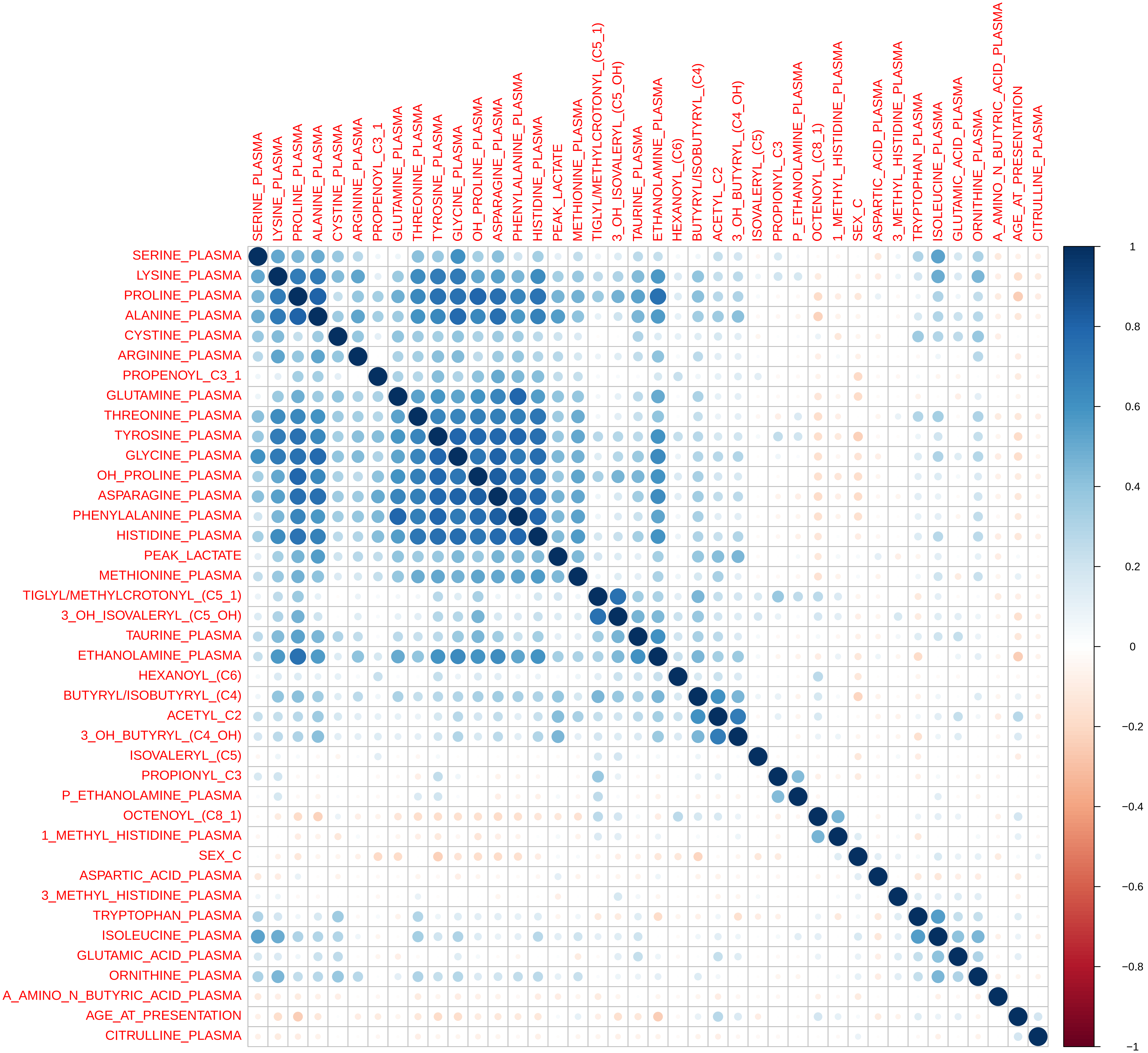

### S Fig 2

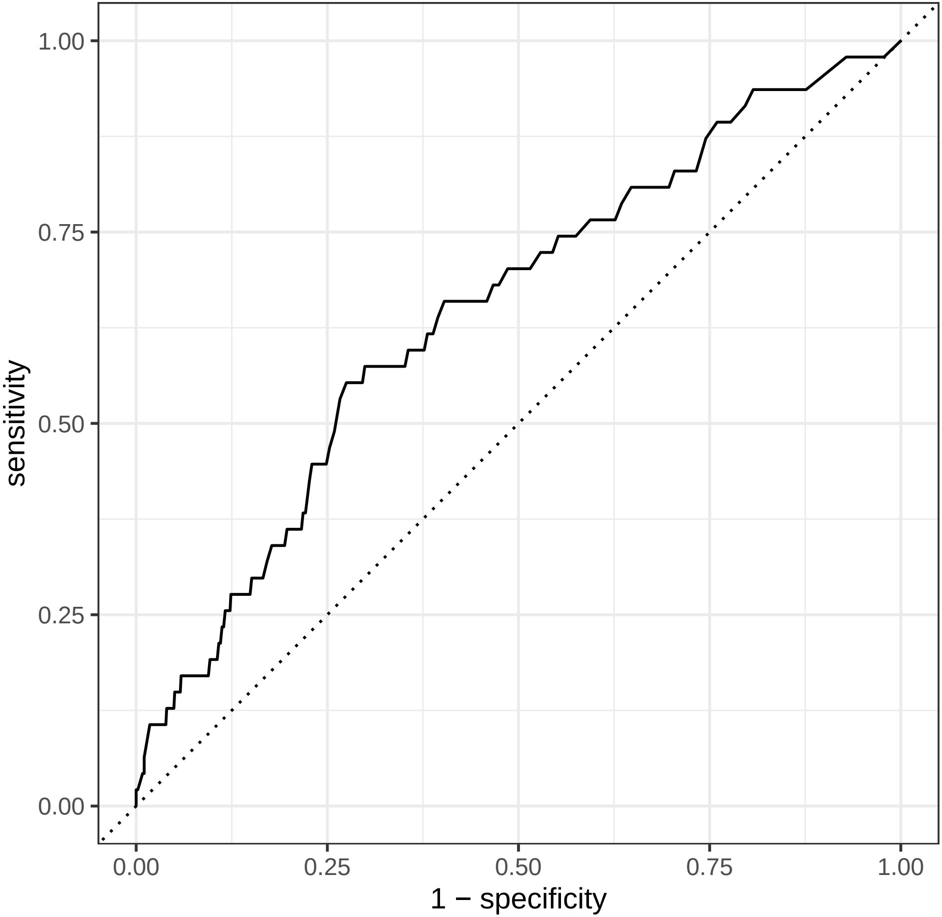

### S Fig 2

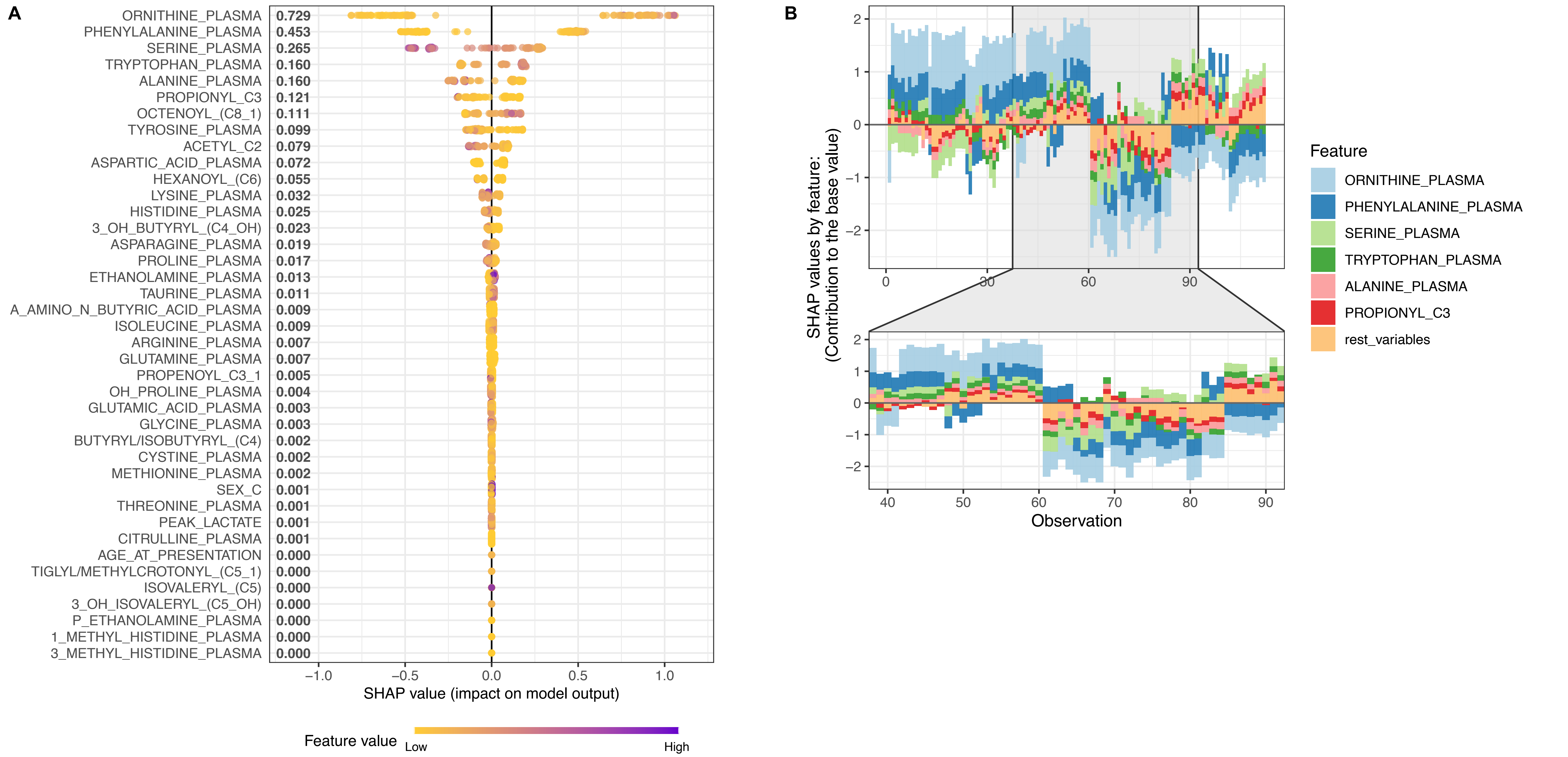

### S Fig 4

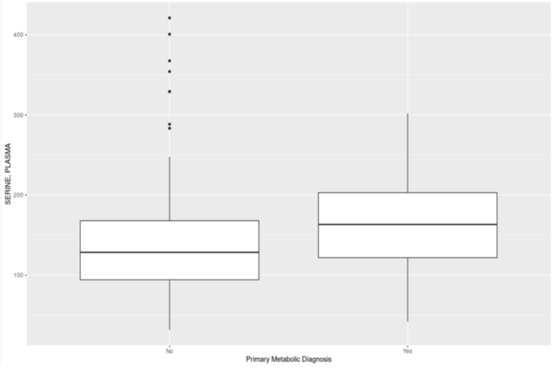
