## Supplementary material for "Machine Learning Improves the Predictive Utility of Lactic Acid in Hospitalized Infants": S Table 1

### Supplementary tables

Table 1: Cross-validation Results

| Method | Accuracy | ROC-AUC | $F_{meas}$ | PR-AUC |
|---|---|---|---|---|
| RF untuned | 0.816 | 0.796 | 0.895 | 0.935 |
| RF tuned | 0.832 | 0.833 | 0.904 | 0.945 |
| XGB untuned | 0.859 | 0.831 | 0.913 | 0.944 |
| XGB tuned | 0.824 | 0.863 | 0.897 | 0.962 |

Table 2: Test Set Predictions

| Method | Accuracy | ROC-AUC | $F_{meas}$ | PR-AUC |
|---|---|---|---|---|
| XGB tuned | 0.895 | 0.808 | 0.938 | 0.937 |
